# Retrospective study on the clinicogenomic characteristics of *EGFR* mutant and wildtype NSCLC

**DOI:** 10.1101/2020.08.07.20170027

**Authors:** Chirag Dhar

## Abstract

**Background:** Lung cancer is among the leading causes of mortality. Nearly 90% of all lung cancers are histologically classified as non-small cell lung cancer (NSCLC). A subset of these tumors harbor mutations on the epidermal growth factor receptor gene (*EGFR*) and such patients are candidates for targeted therapy with *EGFR* tyrosine kinase inhibitors (EGFR TKIs).

**Aim:** To compare and contrast the clinicogenomic characteristics of *EGFR* mutant and wildtype NSCLC.

**Methods and results:** A retrospective cohort study design was used to analyze publicly available data on cBioPortal.org. Patients with *EGFR* mutations were more likely to be female, of Asian ethnicity, never-smokers, and diagnosed with lung adenocarcinoma. Metastasis to the pleura, pleural fluid, and liver were common in patients with *EGFR* mutant NSCLC. On the other hand, lymph node, brain and adrenal gland metastases were more common in patients with other mutations. While the median overall survival was about the same in the two groups, progression free survival was significantly shorter in the *EGFR* mutant group. The mutational landscape was significantly different in the two groups with *EGFR* mutant NSCLCs having a lower mutational burden. Differences in copy number alterations between the two groups were also noted.

**Conclusions:** The clinicogenomic profiles of *EGFR* mutant and *EGFR* wildtype significantly differ. Further studies on these differences and underlying mechanisms are likely to lead to new “druggable” targets that overcome EGFR TKI resistance.

## Introduction

Neoplasms of the lung are one of the leading causes of cancer in both men and women and are a major cause of mortality in the American population with an anticipated 135,000 deaths in 2020 (1). To put this in perspective, this number is only slightly less than the number of lives lost to the COVID pandemic in the USA as of August 6, 2020. 80% of all lung cancers are further categorized as non-small cell lung cancer (NSCLC) (2). Major histological subtypes of NSCLC include lung adenocarcinoma, squamous cell carcinoma large cell carcinoma, adenosquamous cell carcinoma, and sarcomatoid carcinoma (3). Recent advances in the clinicogenomics of lung cancer have uncovered the role of epidermal growth factor receptor (*EGFR*) mutations in a significant proportion of NSCLC patients (4)(5). *EGFR* is a member of the receptor tyrosine kinase family and plays an important role in cellular growth, proliferation and signaling. Certain somatic *EGFR* mutations observed in a subset of NSCLC patients cause overamplification leading to constant activation and uncontrolled cell division (3). Exons 18, 19, 20 and 21 of the *EGFR* gene are often the site of these mutations (6)(7)(8). Some of the commonest *EGFR* mutations include inframe deletions of exon 19 and the exon 21 L858R point mutation (3). These mutations are present in nearly a third of all lung adenocarcinomas and predict efficacy to *EGFR* tyrosine kinase inhibitors (TKIs) such as gefitinib and erlotinib(9)(10). Patients receiving TKIs have improved clinical outcomes as compared to those patients that receive conventional chemotherapy (11)(12)(6)(13).

The establishment of cBioPortal.org(14)(15), a central resource for patient and sample level clinicogenomic data in cancer, allows for in-depth analyses and comparisons of various cancer subtypes. In this retrospective study, the clinicogenomic characteristics of EGFR mutant and EGFR wildtype NSCLC were compared and contrasted.

## Study design and methodology

The following schema describes the methods of data collection and analysis in this retrospective cohort study. References to studies utilized are provided here (16)(17)(18)(19)(20)(21)(22)(23)(24)(25)(26)(27)(28).

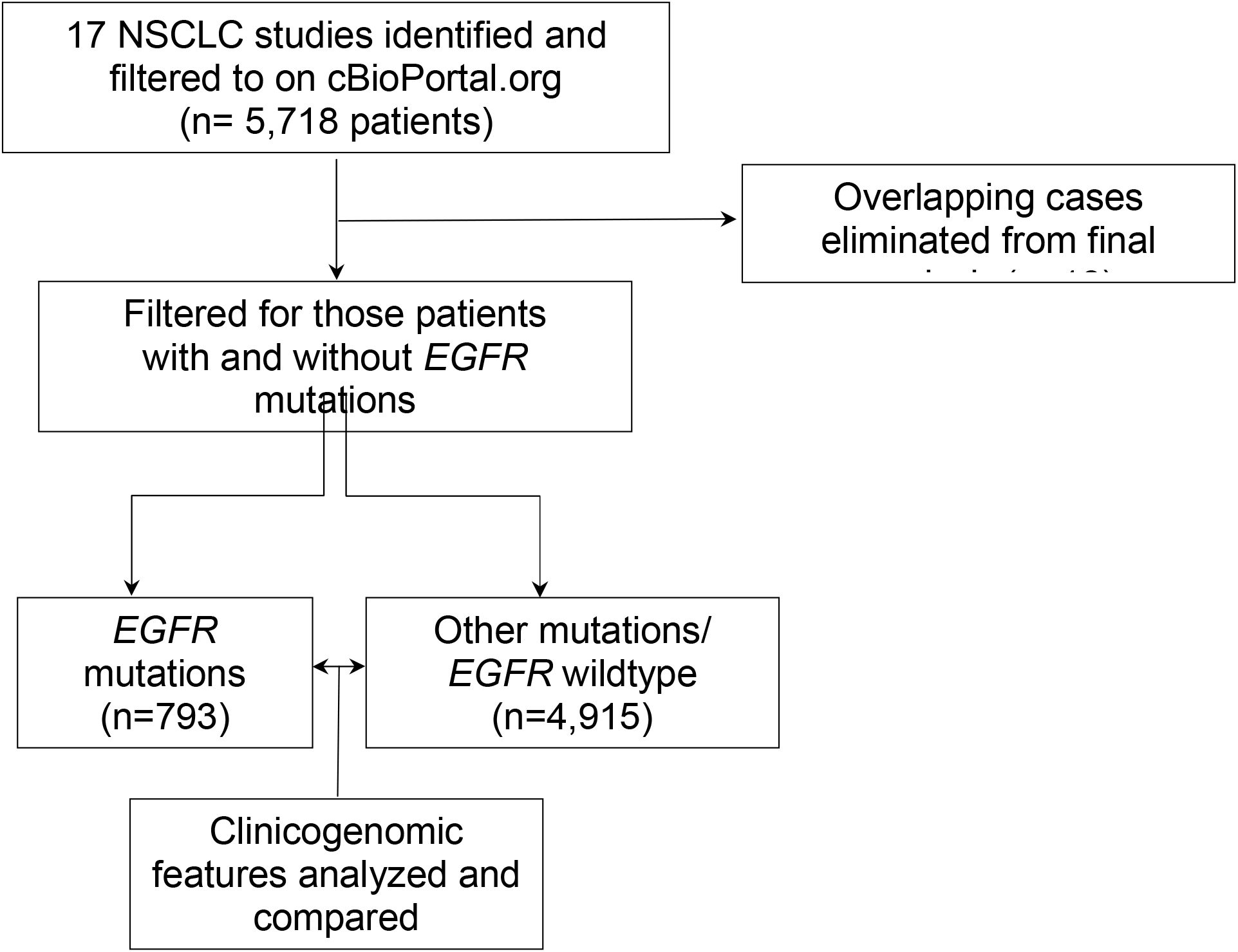

Briefly, 17 cBioPortal.org NSCLC studies representing 5,718 patients were identified and filtered to on September 10^th^, 2020. Of these, 803 were categorized to have *EGFR* mutations while the remaining 4,925 were not. A further analysis on the patient overlap function of cBioPortal revealed 10 cases that were present in both groups. These 10 patients possibly represent multiple samples that had different mutational patterns. For the purpose of this study, these overlapping samples were not analyzed. Subsequently, clinicogenomic features of the two subgroups were compared utilizing the various functions available on cBioPortal. These analyses included a comparision of survival, staging, mutations and copy number variations. Appropriate statistical tests/graphical representations as run by cBioPortal are included in the results. P- and Q-values where appropriate are mentioned. The phrase “*EGFR* wildtype” and “other mutations” are used interchangeably to represent the sub-group that did not harbor *EGFR* mutations.

## Results

### *EGFR* mutations were more common in women, patients of Asian ethnicity, never smokers and those with lung adenocarcinoma

Clinical characteristics of patients with *EGFR* mutations and those with other mutations (*EGFR* wildtype) were compared and are described in detail in table 1. The median age of diagnosis was slightly lower in patients with *EGFR* mutations. These patients were more likely to be never-smokers, women, of Asian ethnicity and diagnosed with lung adenocarcinoma. These findings are in concert with those of other studies (5)(10)(29)(30)(31) on *EGFR* mutant NSCLC and suggest this cohort of more than five thousand patients is likely representative of differences seen in individual studies.

**Table 1:** Clinical characteristics of patients in the two subgroups. Median age in *EGFR* group was 64 years (Range: 36-92, 25^th^ percentile- 57, 75^th^ percentile- 71) while in the other mutations group it was 67 years (Range: 38-93, 25^th^ percentile- 59, 75^th^ percentile- 73) with an absolute p-value by Kruskal Wallis test being 6.25 × 10^−10^ and q-value of 6.07 × 10^−9^. A chi-squared statistic of 155.708 was obtained for the comparison of sex distribution in the two groups (p-value was <10^−6^). Ethnicity data was available for 280 and 2047 patients in the two groups with a significantly higher percentage being of Asian/Chinese ethnicity in the *EGFR* group (Chi-squared test statistic was 397.8601, p-value <10^−6^). Smoking histories of 403 and 2221 patients was available in the two subgroups with never-smokers being significantly higher in the *EGFR* group (Chi-squared test statistic was 296.4889, p-value <10^−6^). 744/805 *EGFR* cases and 3096/4915 other cases were lung adenocarcinoma (Chi-squared test statistic was 2755511, p-value <10^−6^) *all p-values reported to third-decimal place in table

| Clinical characteristic | *EGFR* | Other mutations | P value |
| --- | --- | --- | --- |
| Total number of subjects (%) | 793 (15.8) | 4195 (84.2) | - |
| Median age at diagnosis in years | 64 | 67 | <0.001 |
| Female sex (%) | 64.5 | 41.4 | <0.001 |
| Asian/Chinese ethnicity (%) | 48.1 | 7.8 | <0.001 |
| Never smokers (%) | 38.9 | 7.9 | <0.001 |
| Lung adenocarcinoma (%) | 93.8 | 62.9 | <0.001 |

### *EGFR* mutant cancers were more likely to metastasize than cancers with other mutations

Figure 1 shows the differences in TNM and American Joint Committee on Cancer Code staging between *EGFR* mutant NSCLC and tumors with other mutations. *EGFR* mutant cancers were more likely to involve lymph nodes and to metastasize. Additionally, *EGFR* mutant tumors were more likely to be diagnosed as stage II, III and IV as compared to tumors with other mutations.

**Figure 1:**
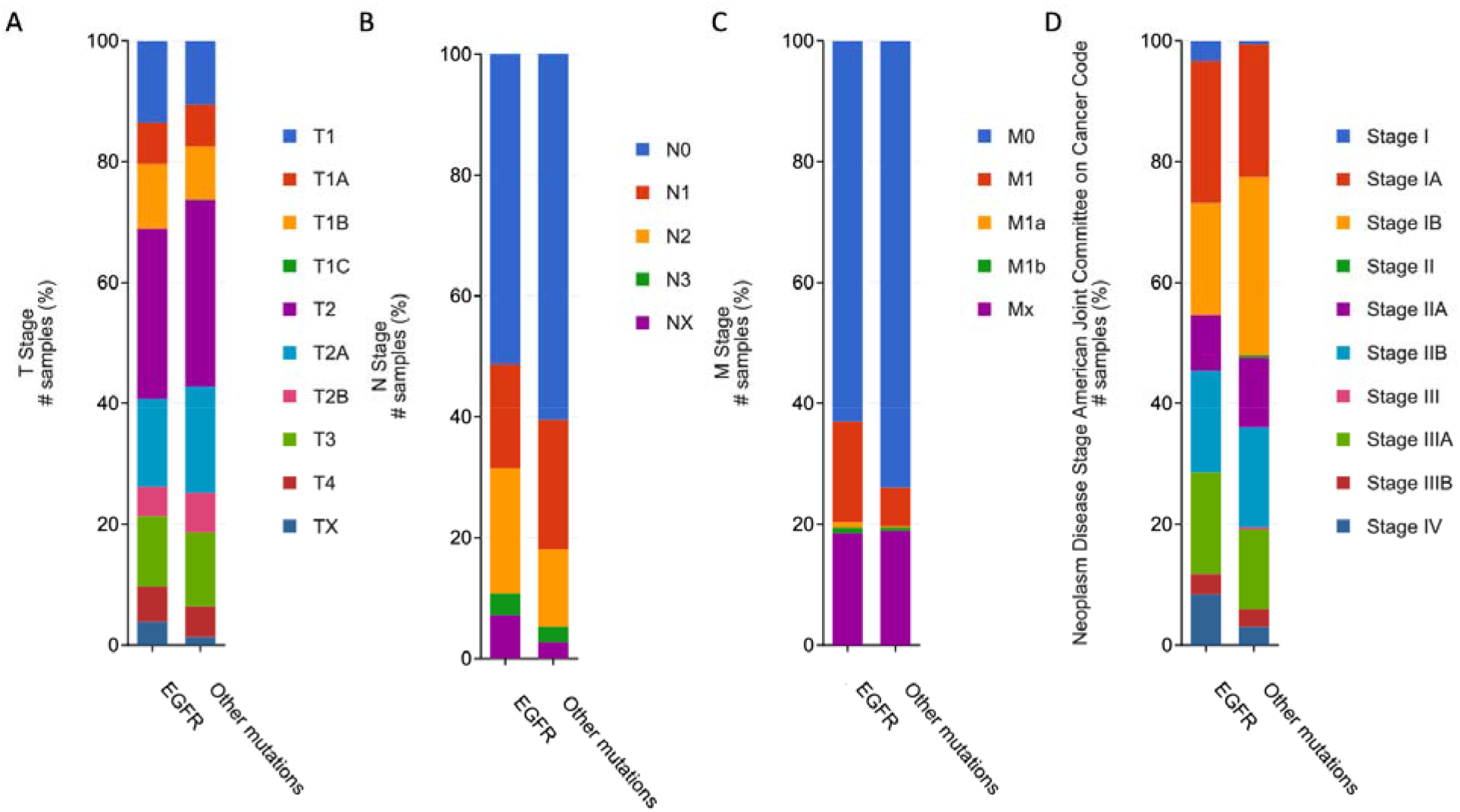
Graphical representation of differences in staging between *EGFR* mutant NSCLC and tumors with other mutations. **A) Differences in T stage**. Non-significant difference (Chi-squared test p-value 0.393) **B) Differences in N stage**. Non-significant difference (Chi-squared test p-value 0.027) **C) Differences in M stage**. Nonsignificant difference (Chi-squared test p-value 0.254) **D) Differences in American Joint Committee on Cancer Code staging**. Significant difference with Chi-squared test p-value of 0.00074

**Metastasis to pleura, pleural fluid and liver were common in patients with *EGFR* mutant NSCLC while metastasis to the lymph node, adrenal gland and brain were more commonly associated with other mutations**. Metastatic samples collected were used as a proxy to indicate frequency and sites of metastases. Metastatic samples were more frequently collected in patients with *EGFR* mutations as indicated by figure 2A. Collected metastatic samples from pleura, pleural fluid and liver were common in patients with *EGFR* mutant NSCLC while metastatic samples from lymph nodes, adrenal gland and brain were common in patients with other mutations. These findings suggest a difference where *EGFR* mutant cancers are likely to metastasize.

**Figure 2:**
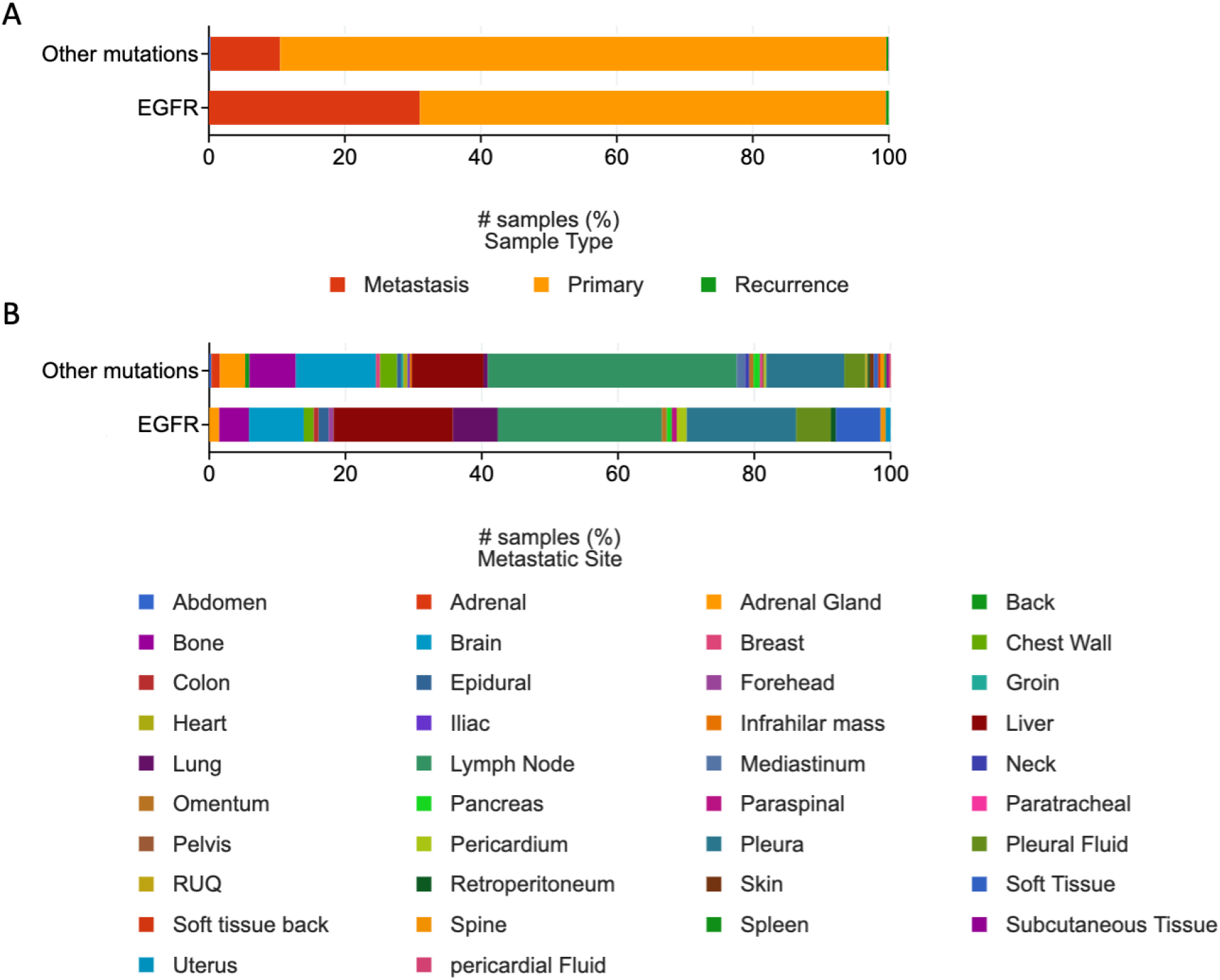
A) Frequency of metastatic sample collection B) Frequency of metastatic sample collection by site. Significant difference between the two groups (Chi-squared test p-value 0.00641)

### Overall survival (OS) is comparable in the two groups while progression free survival (PFS) is shorter in patients with *EGFR* mutations

OS and PFS was compared in the two groups and Kaplan-Meir curves were generated (figure 3A and B). The OS in both the groups were comparable with *EGFR* mutant cancer patients having a median survival of ~49 months and those with other mutations surviving ~53 months (figure 3C). The PFS in patients with EGFR mutations was significantly shorter than those with other mutations (~16 months versus ~30 months, figure 3C).

**Figure 3:**
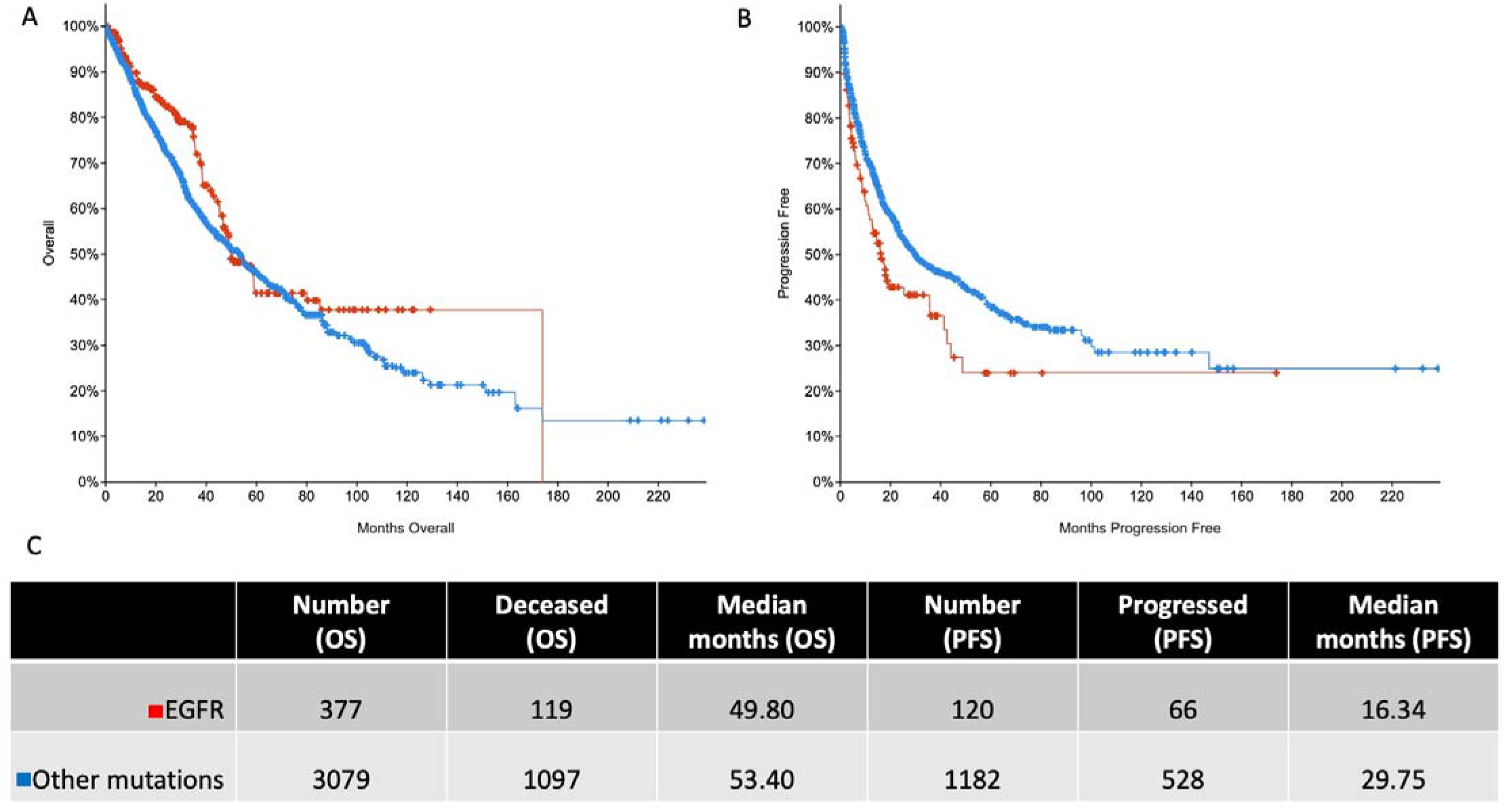
**A) Overall survival (OS) Kaplan-Meir curve**. Logrank test p-value 0.0240 **B) Progression free survival (PFS) Kaplan-Meir curve**. Logrank test p-value 0.0042 **C) Table depicting number of patients where data was available, number that either died or progressed and median survival in months**.

### Patients with other mutations have a higher tumor mutation burden and have higher levels of cell-free DNA

The quantity of cell-free DNA isolated from patients with mutations other than *EGFR* was higher than those with *EGFR* mutations (figure 4A). Similarly, the tumor mutation burden was higher in patients with other mutations (figure 4B).

**Figure 4:**
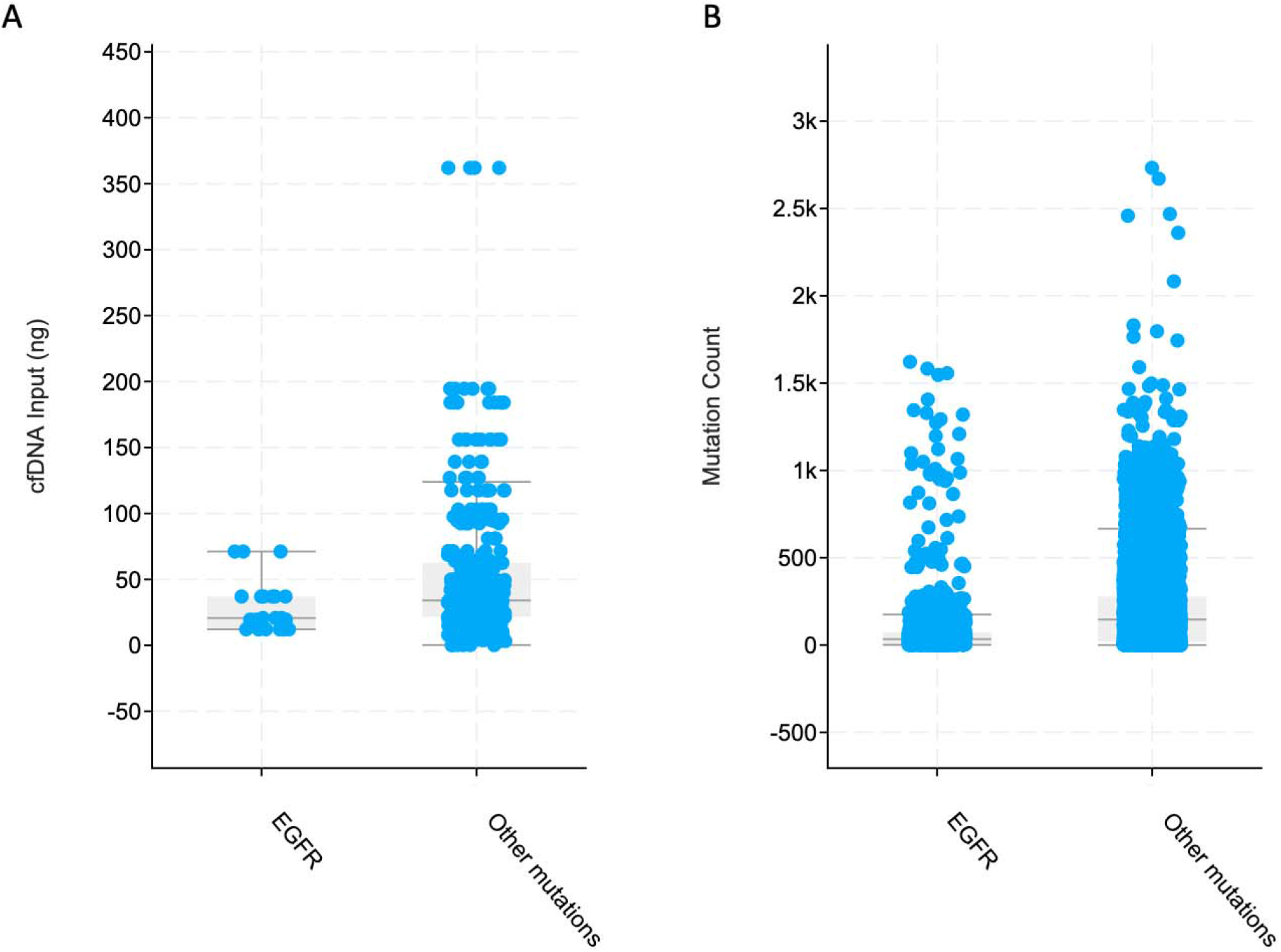
**A) Differences in quantity of cell-free DNA obtained from patients in the two groups.** Nonsignificant difference with median quantities being 20.7 ng and 34 ng in *EGFR* and other mutations group respectively (Chi-squared p-value 0.199) **B) Differences in tumor mutation count**. Median mutation count of 35 in *EGFR* groups versus 151 in other mutations group (Significant difference with Kruskal Wallis test p-value <10^−10^)

### Co-occurrence of mutations and copy number alterations differ in the two groups

Commonly co-occurring mutations in the two groups were analyzed. TP53 mutations were amongst the commonest mutations in both groups and were excluded in this analysis. Similarly, as anticipated *EGFR* mutations were most frequent in the *EGFR* mutant group but have been excluded in the analysis. The most frequently co-occurring mutations in the *EGFR* mutant group (with low frequency in the other group) were in *SAP30L, DEFB4A, IL34, LRRC29, SPINK9, TTC1, REP15, CRIP2, CIA02B, KRTCAP2, REX05, SRP9, TNFRSF12A, CCL2, SH2D1B, AREG, HIST1H3F, TTC31, MRPL10* and *SIAH1*. Commonest mutations in patients with mutations other than *EGFR* were *KRAS, RYR2, MUC16, CSMD3, USH2A, ZFHX4, KEAP1, SYNE1, STK11, NAV3, FLG, SPTA1, FAM135B, XIRP2, FAT3, RYR3, ZNF804A, KMT2C, CUBN* and *Si*. Frequency of occurrence of these mutations are depicted in figure 5A and B.

**Figure 5:**
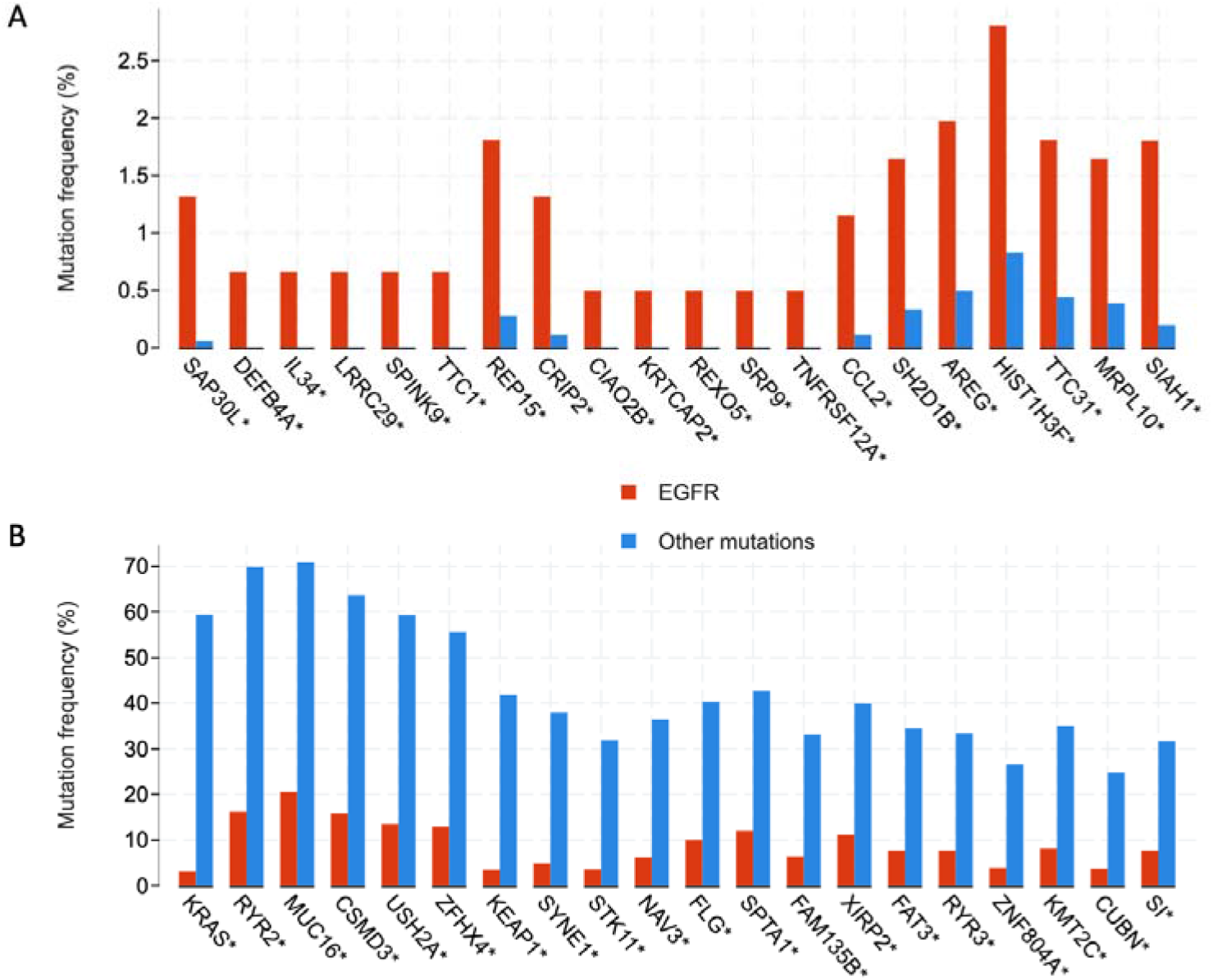
A) and B) Frequency of commonest co-occurring mutations. *asterisked genes represent significant differences.

The commonest copy number alterations were similarly analyzed. As was done with the mutations, *EGFR* amplifications have not been discussed as these are commonly seen in patients with *EGFR* mutations. The commonest amplifications in the *EGFR* group were on *LOC650226, HPVC1, LOC100130849, DKFZP434L 192, CCT6A, SNORA15, SUMF2, VSTM2A-OT1, VOPP1* and *PHKG1* (figure 6A).

The commonest alterations in the other group were amplifications of *DCUN1D1, ATP11B, MCCC1, SOX2, B3GNT5, MCF2L2, LA MP3, KLHL24* and *YEATS2* (figure 6B).

**Figure 6:**
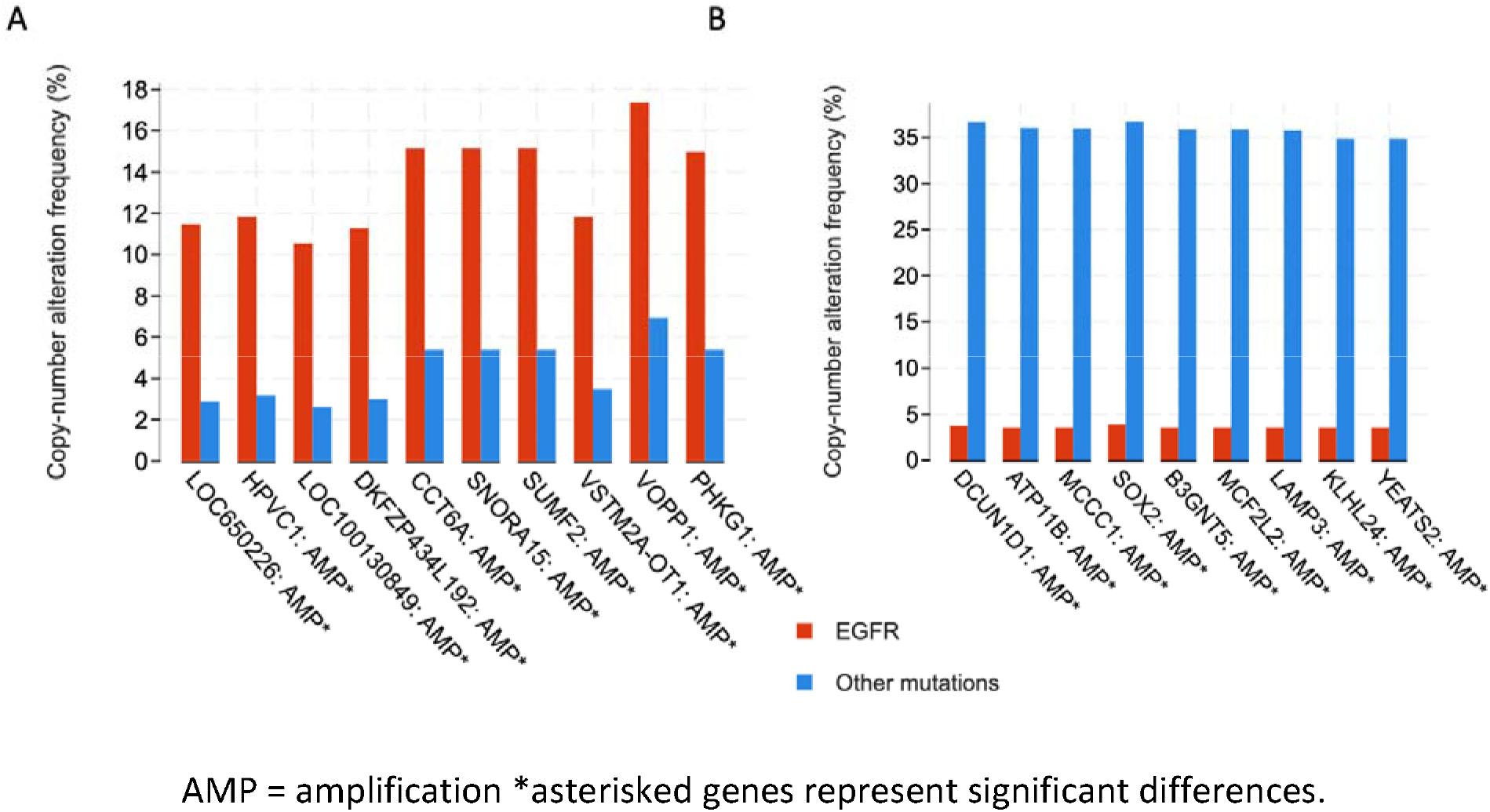
A) and B) Frequency of various copy number alterations in the two groups. AMP = amplification *asterisked genes represent significant differences.

## Discussion

This retrospective study comparing *EGFR* mutant and *EGFR* wildtype NSCLC confirm the commonest clinical characteristics described in other studies. Patients with *EGFR* mutations were more likely to be female, never-smokers and of Asian ethnicity. Lung adenocarcinoma was the commonest histological subtype in these patients. These findings being similar to other studies (5)(10)(29)(30)(31) pointed towards these metadata being representative of differences seen in multiple individual studies. Further, I showed that there were differences in the staging of cancer between the two groups with *EGFR* mutant cancer being more likely to be diagnosed at a more advanced stage. Metastatic samples collected from the two groups suggest that *EGFR* mutant cancer are more likely to metastasize to the pleura and pleural fluid as well as to the liver. On the other hand, NSCLC tumors harboring other mutations were more likely to metastasize to lymph nodes, to the brain and to the adrenal glands. Likely due to these differences in staging and higher likelihood of metastasis in the *EGFR* mutant group, the progression free survival was shorter in patients with *EGFR* mutations. Yet, the difference in median overall survival was only 1 month. These findings are striking in light of targeted therapies for *EGFR* mutant cancers with EGFR TKIs and are strongly suggestive of rapid development of resistance to these therapies (32)(33).

At the genomic level, the tumor mutation burden was higher in *EGFR* wildtype patients. This finding is anticipated as these cancers may lack specific driver mutations and instead rely on multiple mutations for their transformation from normal tissue to cancer. The quantity of cell free DNA though, not significantly different in the two groups, was lower in patients with *EGFR* mutations. The mutation and copy number alteration landscape were also different in the two groups and the commonest genes that were mutated or amplified are listed in the results section.

This study is limited by its retrospective design and therefore is likely to have a degree of convenience sampling. The effects of this error were likely minimal as the clinical characteristics of the two groups were similar to that seen in other prospective studies. Additionally, there were limitations in the quality of data. For example, 6 of 17 included studies did not provide copy-number variation data for each. It is also possible that endpoints such as progression free survival were calculated differently in the various studies included in these analyses. Despite filtering to NSCLC studies, a small number of lung cancers other than NSCLC (<0.5%) also crept through the filters. This number was very small and likely did not alter the results significantly. cBioPortal data does not include specific treatment data and it is possible that a bias towards the null may have occurred in the overall survival data. Other studies have shown better outcomes in patients with *EGFR* mutations when treated with EGFR TKIs (11)(12)(6)(13). For the convenience of this study, NSCLC was broadly divided in to two groups, one for patients with *EGFR* mutations and the other for patients with wildtype *EGFR*. The second group is an oversimplification and includes a gamut of different mutations including some driver mutations such as *KRAS*. Finally, despite showing clear differences in the mutation and copy number alteration landscape in the two groups, this study does not clearly define differences in the molecular mechanisms.

## Conclusions

Such studies on the clinicogenomic features of NSCLC and other cancers are likely to throw light on possible new “druggable” targets. Additionally, hypotheses may be drawn from these studies and taken back to the “bench” to understand specific molecular mechanisms such as resistance to EGFR TKIs (34) or the role of cell-free DNA (35). Future prospective studies and clinical trials are likely to include genomic level and transcriptomic analyses to draw broader clinicogenomic conclusions and lead to significant advances in the management of NSCLC.

## Data Availability

All necessary data are included in the manuscript.

## Notes

### Competing Interest Statement

The authors have declared no competing interest.

### Funding Statement

CD recieced a CREST M.A.S scholarship from ACTRI.

### Summary of Updates

General typos/language improvement. Statistical test results. Expanded methodology

